# Comparative analysis of retracted pre-print and peer-reviewed articles on COVID-19

**DOI:** 10.1101/2022.07.12.22277529

**Authors:** Manraj Singh Sra, Mehak Arora, Archisman Mazumder, Ritik Mahaveer Goyal, Giridara G Parameswaran, Jitendra Kumar Meena

## Abstract

**Introduction:** Due to the accelerated pace and quantum of scientific publication during the COVID-19 pandemic, a large number of articles on COVID-19 have been retracted. Pre-prints though not peer-reviewed offer the advantage of rapid dissemination of new findings. In this study, we aim to systematically compare the article characteristics, time to retraction, social media attention, citations, and reasons for retraction between retracted pre-print and peer-reviewed articles on COVID-19.

**Methods:** We utilized the Retraction Watch database to identify retracted articles on COVID-19 published from 1^st^ January 2020 to 10^th^ March 2022. The articles were reviewed and metadata such as article characteristics (type, category), time to retraction, reasons for retraction, and Altmetric Attention Score (AAS) and citation count were collected.

**Results:** We identified 40 retracted pre-prints and 143 retracted peer-reviewed articles. The median (IQR) retraction time for pre-print and peer-reviewed articles was 29 (10-81.5) days and 139 (63-202) days (p = 0.0001). Pre-prints and peer-reviewed article had median (IQR) AAS of 26.5 (4-1155) and 8 (1-38.5), respectively (p = 0.0082). The median (IQR) citation count for pre-prints and peer-reviewed articles was 3 (0-14) and 3 (0-17), respectively (p = 0.5633). The AAS and citation counts were correlated for both pre-prints (r = 0.5200, p = 0.0006) and peer-reviewed articles(r = 0.5909, p = 0.0001). The commonest reason for retraction for pre-prints and peer-reviewed articles concerns about data and results.

**Conclusion:** The increased adoption of pre-prints results in faster identification of erroneous articles compared to the traditional peer-review process.

## Introduction

Since the start of the COVID-19 pandemic, a large number of articles on epidemiology, diagnostics, clinical management, and other aspects of the disease have flooded the scientific publication space creating doubts over the authenticity and scientific rigor. (Boschiero et al., 2021) It is estimated that 4% of the world’s research output in 2020 on the Dimensions database was COVID-19 related. (Nicholas Fraser & Bianca Kramer, 2021) Due to the growing doubts about research quality, the publication industry put forth retractions of a large number of COVID-19-related articles, although the proportion of articles retracted was nearly the same as other research. (Peterson et al., 2022)

Pre-publication review systems, although effective and efficient, might fail to prevent the publication of invalid literature. There have been instances where retractions have occurred even from high-impact journals and such retractions have happened over months while the typical pre-pandemic delay in retraction was around 3 years. (Else, 2020). During the pandemic, there was a large increase in the number of studies released as pre-prints. Pre-print though not peer-reviewed has the advantage of facilitating rapid dissemination of the findings, but this practice also has the danger of promoting the quick propagation of faulty research. (King, 2020)

Previously descriptive studies on retracted articles related to COVID-19 have been carried out, but comprehensive comparison between pre-print and peer-reviewed articles has not been done. (Shimray, 2021) In this study, we aim to compare the article characteristics, time to retraction, social media attention, citations, and reasons for retraction of peer-print and peer-reviewed articles on COVID-19.

## Materials and methods

### Data Acquisition

In this study, we utilized the Retraction Watch Database maintained by the Center of Scientific Integrity to identify retracted articles on COVID-19/SARS-COV2. (*Retraction Watch Database*, n.d.) This database was accessed on 10^th^ March 2022 under a data use agreement. Using the PubMed ID or DOI of the retracted articles we determined the Altmetric Attention Scores (AAS) and citations on 24^th^ March 2022 from the Altmetric and Dimensions database, respectively. (*Dimensions*, n.d.; *Discover the Attention Surrounding Your Research – Altmetric*, n.d.)

### Search Strategy

Retracted studies on COVID-19/SARS-COV2 were identified by searching for the keywords “COVID-19”, “COVID”, “SARS-COV-2”, “2019-nCOV” and “Coronavirus” in the title of all pre-prints and peer-reviewed articles in the Retraction Watch Database published from 1^st^ January 2020 to 10^th^ March 2022.

### Variables

For our analysis, we collected the name of the journal/pre-print server, country of the first author, article type, reason of retraction, date of publication, date of retraction, AAS, and citations received. Articles were classified as pre-prints if they were ever published on any of the following preprint platforms: medrRxiv, bioRxiv, arXiv, Social Science Research Network (SSRN), OSF Preprints, and Research Square. Articles that were published in peer-reviewed journals and ever never posted on a pre-print platform were classified as peer-reviewed articles. Articles were classified into case reports, clinical studies, commentary/editorials, conference abstracts, letters, meta-analyses, research articles, and review articles. Article types and reasons for retraction were based on the classification used in the Retraction Watch Database. (Supplementary Table 1 and 2). AAS was considered to be zero for all studies with no Altmetric records, as assumed in previous studies. (Serghiou et al., 2021) We also examined if notice of retraction had been displayed on the pre-print platforms and if the full text of the retracted pre-prints was still available.

### Statistical Analysis

All data were entered into Microsoft Office Excel and statistical analysis was done in R. (*R Core Team (2021). R: A Language and Environment for Statistical Computing. R Foundation for Statistical Computing, Vienna, Austria. URL https://www.R-Project.Org/*., n.d.) Categorical variables were summarized as counts and proportions. Continuous variables were checked for normal distribution using the Shapiro-Wilk test. Continuous variables were summarized as mean (standard deviation) or median (inter-quartile range), based on the results from the Shapiro-Wilk test. Categorical variables were compared using Fisher’s exact test and continuous variables were compared using paired t-test or Kruskal-Wallis rank-sum test. Log-rank test was used to compare the time to retraction between pre-print and peer-reviewed articles. All p-values were two-tailed and p-values <0.05 were considered to be significant. Spearman rank correlation test was used to examine the correlation between continuous variables. UpSet plot was created using the UpSetR package to show the intersection of sets for the reason of retraction. (Conway et al., 2017).

## Results

We included 183 articles in the final analysis, out of which 40 were pre-prints and 143 were peer-reviewed articles. (Supplementary Figure 1) Research articles accounted for 62.8% (n=115) of all retracted articles. Among peer-reviewed articles, 6 journals accounted for 34.7% of the retractions. Most of the retracted pre-prints were posted on medRxiv (n=23, 57.5%) followed by the SSRN (n=8, 20%) and bioRxiv (n=7, 17.5%). Characteristics of studies included in the analysis have been summarized in Table 1. All pre-print servers displayed a notice of retraction for the pre-prints included in this study. The full text was available for 77.5% (n=31) of the articles posted, these articles had been posted on medRxiv, bioRxiv, or Research Square. The full text was unavailable for articles posted on SSRN or OSF Preprints.

**Table 1:**
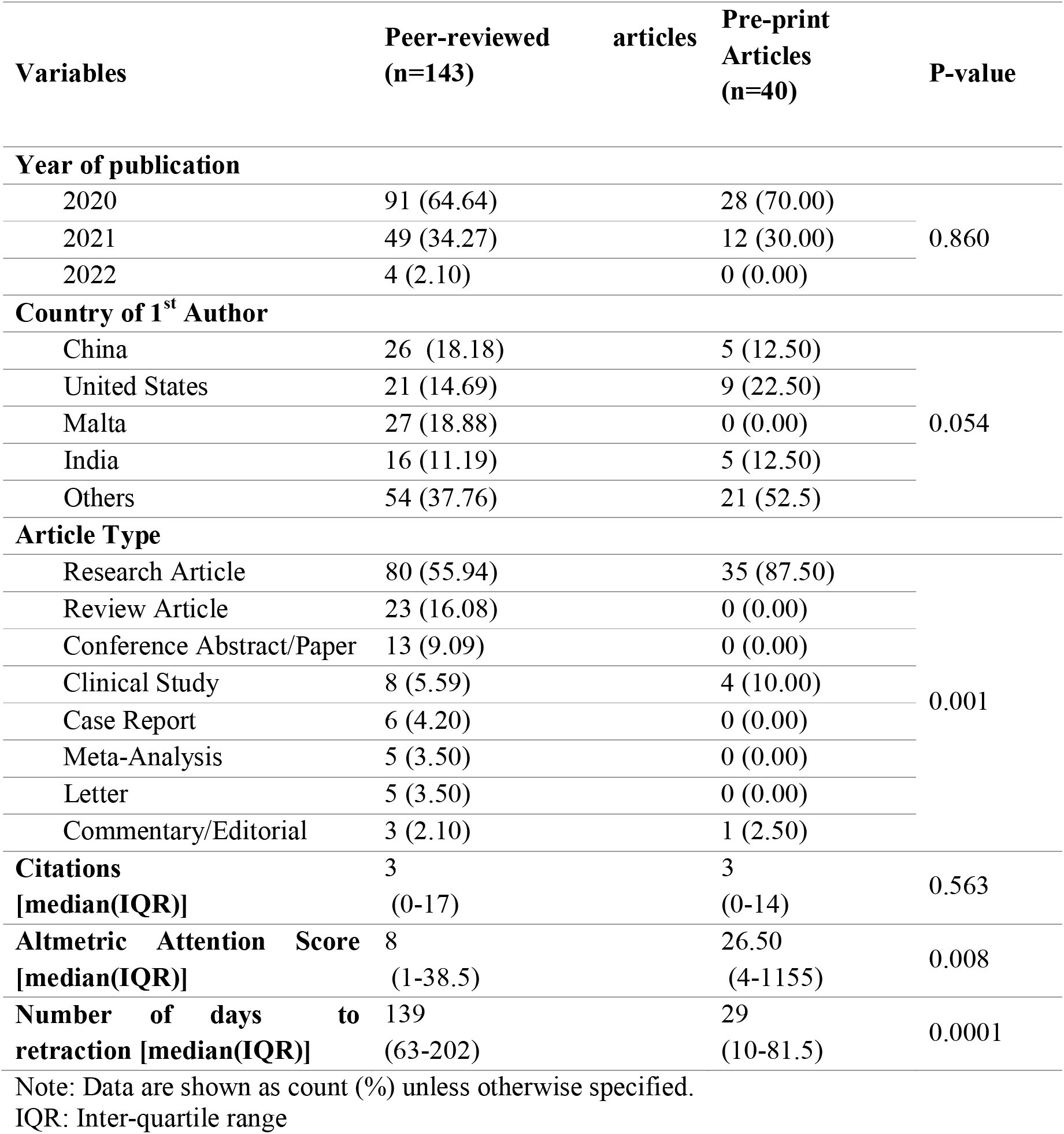
Characteristics of retracted articles

The time to retraction of pre-print article was significantly shorter than peer-reviewed article. (Figure 1 (A); median (IQR): 29 (10-81.5) days vs 139 (63-202) days, p = 0.0001). For pre-print article there was no correlation between AAS and days to retraction (r = -0.0124, p = 0.9392), citations and days to retraction (r = -0.1797, p =0.2673), but AAS and citations were positively correlated (r = 0.5200, p = 0.0006). Similarly peer reviewed articles also did not have any correlation between AAS and days to retraction (r = -0.0918, p = 0.2756) and between citations and days to retraction (r = 0.9000, p=0.2850). But there was a significant correlation between AAS and citations (r = 0.5909, p = 0.0001).

**Fig 1.**
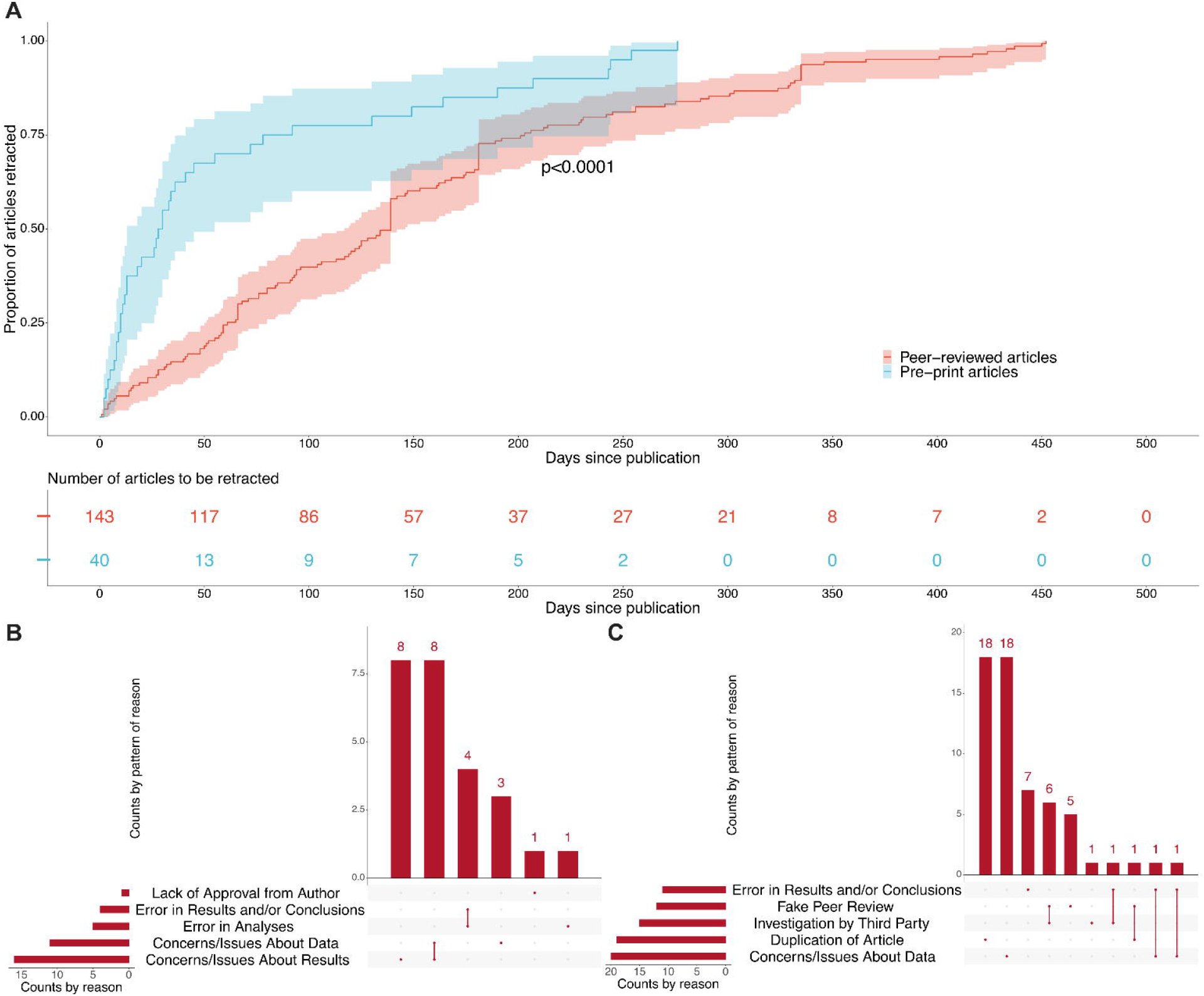
**(A)** Time to retraction of pre-print and peer-reviewed articles. Upset plot showing five commonest reasons for retraction for pre-print **(B)** and peer-reviewed **(C)** publications

Figure 1 (B) and (C) show that concerns/issues about data and results were the commonest cause for retraction of both pre-print as well as peer-reviewed articles. Reasons for retraction of pre-prints and peer-reviewed articles by article type have been detailed in Supplementary Tables 3 and 4, respectively.

## Discussion

The surge in spurious articles on COVID-19 has made it imperative to develop methods for the rapid identification and retraction of such studies. The key finding of our study was that the pre-print articles had a statistically significant smaller delay in retraction compared to peer-reviewed articles. Preprint servers have increased the speed with which scientific information can be submitted, disseminated, and critiqued. Since pre-prints are not paywalled they allow researchers can get valuable feedback from a vast community of peers. (Sarabipour et al., 2019) Even for peer-reviewed articles, previous studies have shown that more retractions happen in open access journals and because of post-publication scrutiny by the scientific community. (Avissar-Whiting, 2022) Additionally, because pre-printed articles are not peer-reviewed, people reading the articles are more critical of the results, methodology, and conclusions which inflates the chances of noticing errors. However, further studies are needed to examine this hypothesis as some have argued that during the pandemic, non-peer-reviewed scientific information was used without caution and scrutiny. (Ravinetto et al., 2021) A study on all biology and medicine-related papers showed that downstream retraction of pre-printed articles is faster. (Avissar-Whiting, 2022) Considering that the quality of the article doesn’t improve much between a preprint and the final peer-reviewed article, it isn’t surprising that earlier and easier access to the results for a critical appraisal can lead to faster retractions. (Carneiro et al., 2020) In our study, we found no correlation between the time to retraction and the AAS and citations. Previously it has been shown that retracted articles receive greater social media attention than similar non-retracted articles. (Serghiou et al., 2021) But non-scientific audiences and bots contribute to a large portion of the social media discussions of retracted articles, thus explaining the lack of a correlation between the time to retraction and the AAS. (Abhari et al., 2022)

Another interesting point to note is the difference in the causes of retraction between preprints and peer-reviewed articles. Some peer-reviewed articles have been retracted for reasons like fake peer review and duplication of articles, which exposes the faults of the peer-review process. A non-peer-reviewed or duplicate article that is vetted as an original peer-reviewed article can spread scientific misinformation, potentially easier than preprints that are mentioned to be non-peer-reviewed. On the other hand, concerns/issues with the results were the most common cause of retraction of preprints.

Our study has some limitations. Firstly this was an exploratory study aimed to compare pre-print and peer-reviewed retractions of COVID-19-related articles. However, our study design and scope are insufficient to conclude causality, and further research is needed to understand the causes leading to a delay in retractions. Also, we did not have longitudinal data of the AAS and citations to compare pre and post-retraction differences. Possibly some retracted articles might have gotten attention after retraction leading to an increase in AAS post retraction.

## Conclusion

The present study shows that there is another benefit to encouraging people to put up their work on preprint servers in the form of earlier recognition of errors and open review leading to prompt retractions. As we go forward, the scientific community should promote preprints as they provide wider and faster reach, more attention, and rapid peer feedback. Importantly, future studies should aim at developing methods for the rapid identification of studies requiring retractions.

## Data Availability

Data used in this is available subject to a standard data use agreement from The Centre for Scientific Integrity, the parent non-profit organization of Retraction Watch.

http://retractiondatabase.org/RetractionSearch.aspx?

## Data availability statement

Data used in this is available subject to a standard data use agreement from The Centre for Scientific Integrity, the parent non-profit organization of Retraction Watch. Web link: http://retractiondatabase.org/RetractionSearch.aspx?

## Contributors

MSS, GGP, and JKM conceptualized the study. MSS, MA, AM, and JKM were involved in data collection and analysis. MSS, MA, AM, and RG wrote the first draft of the manuscript. GGP and JKM provided critical revisions to the manuscript and supervised the study.

