## Supplementary Material for "Comparative analysis of retracted pre-print and peer-reviewed articles on COVID-19"

**Supplementary Table 1:** List of article types

**Supplementary Table 2:** List of reasons for retraction

**Supplementary Figure 1:** Flow chart showing the process of study selection

**Supplementary Table 3:** Reasons for retraction of pre-print articles stratified by type of articles

**Supplementary Table 4:** Reasons for retraction of peer-reviewed articles stratified by type of articles

**Supplementary Table 1:** List of article types

| Article Types | Description |
| --- | --- |
| Case Report | Article describing the signs, symptoms, diagnosis and resolution of a single or very limited number of patients |
| Clinical Study | Research study of humans where the outcome of a group undergoing a specific treatment is compared to a non-treatment group |
| Commentary/Editorial | Opinion piece, either by editor or guest author |
| Conference Abstract/Paper | Item published in conference proceedings |
| Letter | Any type of item published in the Correspondence or Letter (to the Editor) section of a journal |
| Meta-Analysis | An analysis of the combined data from several separate studies |
| Research Article | Published item describing a hypothesis, means of exploring the hypothesis, the results of the exploration, and the conclusions drawn from the results |
| Review Article | Evaluation of literature concerning a topic, without the presentation of new data |

**Supplementary Table 2:** List of article types

| Reason | Description |
| --- | --- |
| Bias Issues or Lack of Balance | Failure to maintain objectivity in the presentation or analysis of information |
| Cites Retracted Work | A retracted item is used in citations or referencing |
| Concerns/Issues About Authorship | Any question, controversy or dispute over the rightful claim to authorship, excluding forged authorship |
| Concerns/Issues About Data | Any question, controversy or dispute over the validity of the data |
| Concerns/Issues about Referencing/Attributions | Any question, controversy or dispute over whether ideas, analyses, text or data are properly credited to the originator |
| Concerns/Issues About Results | Any question, controversy or dispute over the validity of the results |
| Conflict of Interest | Authors having affiliations with companies, associations, or institutions that may serve to influence their belief about their findings |
| Copyright Claims | Dispute concerning right of ownership of a publication |
| Duplication of Article | Also known as “self-plagiarism”.  Used when an entire published item, or undefined sections of it, written by one or all authors of the original article, are repeated in the original article without appropriate citation. |
| Duplicate Publication through Error by Journal/Publisher | Used when a Journal or Publisher incorrectly publishes the same article more than once. Differs from Duplication of Article, which is typically due to dual submission by the article’s authors. |
| Error by Journal/Publisher | A mistake attributed to a Journal Editor or Publisher |
| Error in Analyses | A mistake made in the evaluation of the data or calculations |
| Error in Data | A mistake made in the data, either in data entry, gathering or identification |
| Error in Methods | A mistake made in the experimental protocol, either in following the wrong protocol, or in erring during the performance of the protocol |
| Error in Results and/or Conclusions | A mistake made in determining the results or establishing conclusions from an experiment or analysis |
| Euphemisms for Plagiarism | The notice does not clearly state that the authors reused ideas, text, or images, without suitable citation, from items published by those not the authors |
| Fake Peer Review | The peer review was intentionally not performed in accordance with the journal’s guidelines or ethical standards |
| Investigation by Journal/Publisher | An evaluation of allegations by the Journal or Publisher |
| Investigation by Third Party | An evaluation of allegations by a person, company or institution not the Authors, Journal, Publisher or ORI |
| Lack of Approval from Company/Institution | Failure to obtain agreement from original author(s) |
| Lack of Approval from Third Party | Failure to obtain agreement from original author(s) |
| Lack of IRB/IACUC Approval | Failure to obtain consent from the institutional ethical review board overseeing human or animal experimentation prior to initiation of study, or failure to provide proof of such |
| Lack of Approval from Author | Failure to obtain agreement from original author(s) |
| Plagiarism of Article | Used when an entire published item, or undefined sections of it, and not written by one or all authors of the original article, are repeated in the original article without appropriate citation. |
| Results Not Reproducible | Experiments conducted, using the same materials and methods, that fail to replicate the finding of the original article |
| Retract and Replace | The permanent change of an item to a non-citable status, with a subsequent republication by the same journal after substantial changes to the item |
| Unreliable Data | The accuracy or validity of the data is questionable |
| Unreliable Results | The accuracy or validity of the results is questionable |
| Upgrade/Update of Prior Notice | Either a change to or affirmation of a prior notice |
| Withdrawal | The original article is removed from access on the Journal’s publishing platform. |

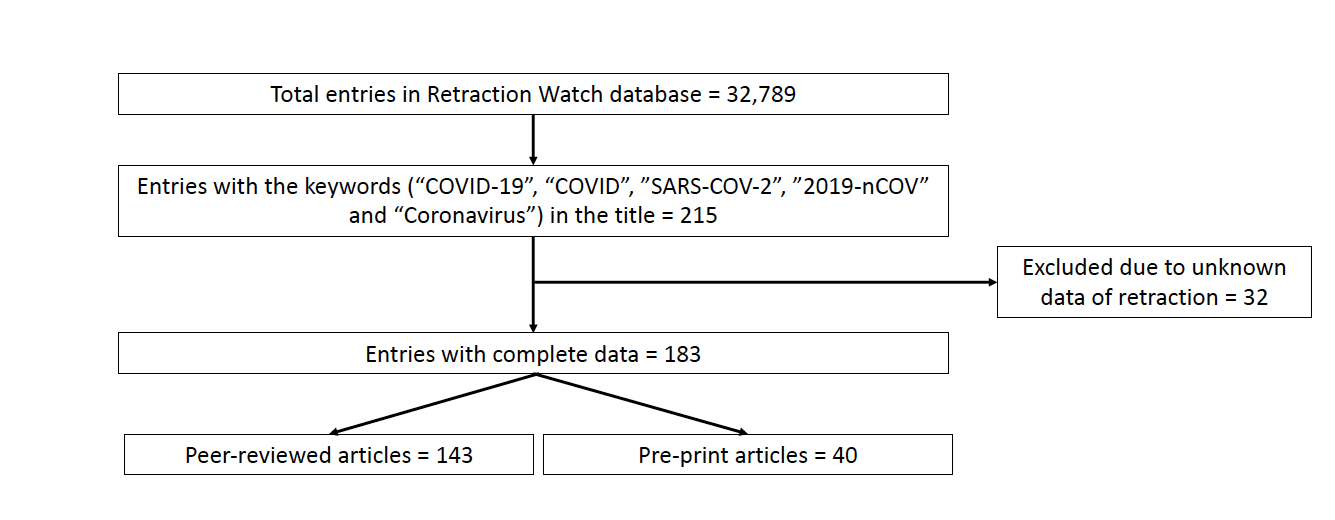
**Supplementary Figure 1:** Flow chart showing process of study selection

**Supplementary Table 3:** Reasons for retraction of pre-print articles stratified by type of articles

| Reason | Count | Percentage |
| --- | --- | --- |
| Clinical Study (n=4) |  |  |
| Concerns/Issues About Data | 1 | 25 |
| Concerns/Issues About Results | 1 | 25 |
| Date of Retraction/Other Unknown | 1 | 25 |
| Error in Analyses | 1 | 25 |
| Error in Data | 1 | 25 |
| Error in Results and/or Conclusions | 1 | 25 |
| Retract and Replace | 1 | 25 |
| Commentary/Editorial (n=1) |  |  |
| Lack of Approval from Third Party | 1 | 100 |
| Research Article (n=35) |  |  |
| Bias Issues or Lack of Balance | 1 | 2.86 |
| Concerns/Issues About Data | 10 | 28.57 |
| Concerns/Issues About Referencing/Attributions | 1 | 2.86 |
| Concerns/Issues About Results | 15 | 42.86 |
| Conflict of Interest | 2 | 5.71 |
| Copyright Claims | 1 | 2.86 |
| Duplication of Article | 1 | 2.86 |
| Error in Analyses | 4 | 11.43 |
| Error in Data | 1 | 2.86 |
| Error in Results and/or Conclusions | 3 | 8.57 |
| Ethical Violations by Author | 1 | 2.86 |
| Informed/Patient Consent - None/Withdrawn | 1 | 2.86 |
| Lack of Approval from Author | 1 | 2.86 |
| Lack of Approval from Third Party | 2 | 5.71 |
| Lack of IRB/IACUC Approval | 2 | 5.71 |
| Objections by Third Party | 1 | 2.86 |
| Unreliable Results | 2 | 5.71 |

Note: One study can have more than one reason for retraction

**Supplementary Table 4:** Reasons for retraction of peer-reviewed articles stratified by type of articles

| Reason | Count | Percentage |
| --- | --- | --- |
| Case Report (n=6) |  |  |
| Concerns/Issues About Data | 1 | 16.67 |
| Concerns/Issues About Results | 1 | 16.67 |
| Copyright Claims | 1 | 16.67 |
| Duplication of Article | 2 | 33.33 |
| Error in Data | 2 | 33.33 |
| Error in Results and/or Conclusions | 2 | 33.33 |
| Fake Peer Review | 1 | 16.67 |
| Clinical Study (n=8) |  |  |
| Concerns/Issues About Data | 2 | 25 |
| Concerns/Issues About Results | 1 | 12.5 |
| Duplicate Publication through Error by Journal/Publisher | 1 | 12.5 |
| Error in Analyses | 1 | 12.5 |
| Error in Methods | 1 | 12.5 |
| Investigation by Journal/Publisher | 1 | 12.5 |
| Lack of Approval from Company/Institution | 1 | 12.5 |
| Lack of IRB/IACUC Approval | 3 | 37.5 |
| Results Not Reproducible | 1 | 12.5 |
| Retract and Replace | 1 | 12.5 |
| Unreliable Data | 1 | 12.5 |
| Unreliable Results | 2 | 25 |
| Upgrade/Update of Prior Notice | 2 | 25 |
| Commentary/Editorial (n=3) |  |  |
| Cites Retracted Work | 1 | 33.33 |
| Concerns/Issues About Referencing/Attributions | 1 | 33.33 |
| Error by Journal/Publisher | 1 | 33.33 |
| Retract and Replace | 1 | 33.33 |
| Conference Abstract/Paper (n=13) |  |  |
| Concerns/Issues About Referencing/Attributions | 6 | 46.15 |
| Duplication of Article | 6 | 46.15 |
| Fake Peer Review | 6 | 46.15 |
| Investigation by Journal/Publisher | 6 | 46.15 |
| Investigation by Third Party | 12 | 92.31 |
| Not Presented at Conference | 1 | 7.69 |
| Letter (n=3) |  |  |
| Concerns/Issues About Authorship | 1 | 33.33 |
| Concerns/Issues About Results | 1 | 33.33 |
| Duplicate Publication through Error by Journal/Publisher | 1 | 33.33 |
| Duplication of Article | 2 | 66.66 |
| Meta-Analysis (n=5) |  |  |
| Cites Retracted Work | 1 | 20.00 |
| Concerns/Issues About Data | 1 | 20.00 |
| Error by Journal/Publisher | 1 | 20.00 |
| Error in Analyses | 1 | 20.00 |
| Error in Data | 1 | 20.00 |
| Error in Methods | 2 | 40.00 |
| Error in Results and/or Conclusions | 1 | 20.00 |
| Investigation by Third Party | 2 | 40.00 |
| Plagiarism of Article | 1 | 20.00 |
| Retract and Replace | 1 | 20.00 |
| Upgrade/Update of Prior Notice | 2 | 40.00 |
| Research Article (n=80) |  |  |
| Breach of Policy by Author | 2 | 2.5 |
| Concerns/Issues About Data | 13 | 16.25 |
| Concerns/Issues About Image | 1 | 1.25 |
| Concerns/Issues About Referencing/Attributions | 2 | 2.5 |
| Concerns/Issues About Results | 5 | 6.25 |
| Concerns/Issues About Third Party Involvement | 1 | 1.25 |
| Conflict of Interest | 3 | 3.75 |
| Copyright Claims | 2 | 2.5 |
| Duplicate Publication through Error by Journal/Publisher | 5 | 6.25 |
| Duplication of Article | 5 | 6.25 |
| Duplication of Image | 2 | 2.5 |
| Error in Analyses | 7 | 8.75 |
| Error in Cell Lines/Tissues | 1 | 1.25 |
| Error in Data | 1 | 1.25 |
| Error in Image | 1 | 1.25 |
| Error in Methods | 5 | 6.25 |
| Error in Results and/or Conclusions | 6 | 7.5 |
| Ethical Violations by Author | 2 | 2.5 |
| Euphemisms for Plagiarism | 2 | 2.5 |
| Fake Peer Review | 5 | 6.25 |
| False Affiliation | 3 | 3.75 |
| FALSE/Forged Authorship | 3 | 3.75 |
| Hoax Paper | 1 | 1.25 |
| Informed/Patient Consent - None/Withdrawn | 1 | 1.25 |
| Investigation by Company/Institution | 2 | 2.5 |
| Investigation by Journal/Publisher | 3 | 3.75 |
| Investigation by Third Party | 1 | 1.25 |
| Lack of Approval from Third Party | 6 | 7.5 |
| Lack of IRB/IACUC Approval | 2 | 2.5 |
| Misconduct by Author | 1 | 1.25 |
| Objections by Author(s) | 1 | 1.25 |
| Objections by Third Party | 1 | 1.25 |
| Paper Mill | 1 | 1.25 |
| Plagiarism of Article | 2 | 2.5 |
| Plagiarism of Data | 2 | 2.5 |
| Retract and Replace | 1 | 1.25 |
| Rogue Editor | 1 | 1.25 |
| Taken via Peer Review | 1 | 1.25 |
| Unreliable Data | 4 | 5 |
| Unreliable Results | 8 | 10 |
| Upgrade/Update of Prior Notice | 5 | 6.25 |
| Review Article (n=23) |  |  |
| Bias Issues or Lack of Balance | 1 | 4.35 |
| Concerns/Issues About Data | 3 | 13.04 |
| Concerns/Issues About Results | 1 | 4.35 |
| Conflict of Interest | 1 | 4.35 |
| Duplication of Article | 4 | 17.39 |
| Error in Analyses | 1 | 4.35 |
| Error in Data | 2 | 8.70 |
| Error in Methods | 1 | 4.35 |
| Error in Results and/or Conclusions | 2 | 8.70 |
| Euphemisms for Plagiarism | 1 | 4.35 |
| Plagiarism of Article | 4 | 17.39 |
| Retract and Replace | 2 | 8.70 |

Note: One study can have more than one reason for retraction
